# Mechanosensitive TRPV4 immunohistochemistry improves deep learning-based classification of ductal carcinoma in situ beyond H&E morphology

**DOI:** 10.64898/2025.12.20.25342730

**Authors:** Janghyun Yoo, Raghav Karthikeyan, Kashi Kamat, Christopher Chan, Shabnam Samankan, Elham Arbzadeh, Arnold Schwartz, Patricia S. Latham, Inhee Chung

## Abstract

Ductal carcinoma in situ (DCIS) spans a biologic continuum from atypical ductal hyperplasia (ADH) to high-grade lesions with variable risk of progression to invasive ductal carcinoma (IDC), yet morphologic assessment by hematoxylin and eosin (H&E) remains diagnostically limited, particularly at the benign versus ADH/low-grade DCIS boundary. TRPV4, a mechanosensitive ion channel with pathology-dependent subcellular localization in DCIS, offers a biologically motivated immunohistochemical (IHC) marker that may refine classification beyond routine H&E assessment. We tested whether deep learning models trained on TRPV4 IHC outperform H&E-based models across the DCIS progression spectrum. We assembled a multi-institutional cohort of paired H&E and TRPV4 IHC whole-slide images from 108 patients, comprising an internal development cohort (n=69), an external test cohort (n=39), yielding 24,248 annotated tiles. Histopathological tiles from annotated regions were grouped into four ordered classes: normal/benign, ADH/low-grade DCIS, high-grade DCIS, and IDC. Xception and EfficientNet-B0 convolutional neural networks were trained with patient-level 3-fold cross-validation on the development cohort and evaluated as ensembles on the external test cohort. On external patient-level testing, H&E ensembles achieved macro-F1 values of 0.43 to 0.44 and macro-AUC values of 0.73 to 0.80, whereas TRPV4 IHC ensembles improved performance to macro-F1 values of 0.68 to 0.72 and macro-AUC values of 0.91 to 0.92, corresponding to a 54.5-67.4% relative improvement in patient-level macro-F1. Per-class analyses showed the largest gains with TRPV4 IHC versus H&E for ADH/low-grade DCIS (AUC, 0.83–0.84 vs 0.70–0.81) and IDC (AUC, 0.74–0.79 vs 0.65–0.66). These findings support TRPV4 IHC as a mechanistically grounded complement to H&E that improves patient-level discrimination across the DCIS progression spectrum, with the strongest gains for ADH/low-grade DCIS and IDC, in a pilot multi-institutional setting.

## Introduction

Ductal carcinoma in situ (DCIS) is a non-invasive breast cancer characterized by the proliferation of neoplastic epithelial cells confined within breast ducts and accounts for approximately 20% of newly diagnosed breast cancer cases (1–3). Fewer than half of DCIS lesions progress to IDC if left untreated (4, 5), yet there is no reliable method to predict which cases will become invasive. Clinicopathologic factors and multigene assays have been developed to predict recurrence risk but show limited ability to predict IDC progression (2, 6–11). As a result, the current standard of care frequently favors surgical intervention, raising concern for overtreatment (1, 3, 6). Histopathologic evaluation of hematoxylin and eosin (H&E)-stained sections remains central to diagnosis and grading (1, 2, 12–15), but morphology-based risk assessment is inherently subjective and can reduce diagnostic accuracy and reproducibility, especially at biologically and clinically important boundaries (16–19).

In routine practice, breast lesions are categorized as benign, atypical ductal hyperplasia (ADH), DCIS, or IDC, with DCIS further stratified into low-, intermediate-, and high-grade based on nuclear atypia, architecture, and necrosis (1, 12–15). However, these categories lie along a biologic continuum rather than representing discrete states. Small differences in nuclear features or necrosis can shift a lesion between ADH and low-grade DCIS or between DCIS grades, making some distinctions particularly fragile. The most diagnostically challenging and clinically consequential transitions include: (1) normal/benign epithelium versus ADH/low-grade DCIS (LG-DCIS), (2) ADH/LG-DCIS versus high-grade DCIS (HG-DCIS), and (3) the distinction of DCIS from IDC (20–22). Among these, the distinction between benign epithelium and ADH/low-grade DCIS is particularly challenging because small morphological differences can alter both diagnosis and clinical management (12, 16, 20, 21). In this study, we use four biologically ordered diagnostic groups along the DCIS-related progression axis to capture the broader spectrum of disease, while placing primary clinical emphasis on the benign versus ADH/low-grade DCIS boundary. Intermediate-grade DCIS cases, which are known to show particularly low interobserver agreement (17–20), were omitted from this initial deep learning analysis to reduce label noise in a modest-sized cohort.

To address limitations of morphology alone, deep learning approaches have been applied to breast pathology to computationally distinguish normal or benign tissue from DCIS, and between DCIS and IDC, typically using morphologic features from H&E (23–31). While these methods may enhance overall classification performance and reproducibility relative to purely manual assessment, they remain constrained by their reliance on morphology and therefore inherit the subjectivity and ambiguity of traditional grading. One promising strategy is to complement morphology with biologically grounded markers that may improve classification across this progression spectrum.

We and others have characterized the mechanosensitive, calcium-permeable ion channel TRPV4 as a regulator of cellular responses to mechanical stress (32–35). In our recent study, we showed that in confined DCIS ducts, ductal cell crowding stress drives TRPV4 relocalization from intracellular compartments to the plasma membrane as part of a crowding-induced pro-invasive mechanotransduction program (35). In our subsequent analytical-validation and proof-of-concept prognostic study, we showed that PM-TRPV4 can be scored reproducibly on human formalin-fixed, paraffin-embedded (FFPE) DCIS sections and serves as a TRPV4 localization-based readout of pro-invasive mechanotransduction rather than total channel abundance (36). Although PM-TRPV4-positive fields were more frequent in higher-grade DCIS, PM-TRPV4 also remained associated with invasive progression independent of histologic grade and estrogen receptor status, with the strongest exploratory clinical signal in lower-grade disease (36). Together, these findings suggest that TRPV4 IHC may encode biologically relevant information beyond routine morphology and therefore motivate direct comparison of deep learning model performance on TRPV4 IHC versus H&E for DCIS classification.

In the present study, we tested whether deep learning models trained on TRPV4 IHC improve DCIS-related classification compared with models trained on H&E alone, particularly at the diagnostically challenging benign versus ADH/low-grade DCIS boundary. Using a multi-institutional cohort with an internal development set from the University of Virginia (UVA) and an independent external test set from George Washington University (GWU), we asked whether TRPV4 IHC improves discrimination at this clinically important boundary while also improving classification across the broader DCIS progression spectrum and generalizing across institutions. Tiles from pathologist-annotated regions were grouped into four ordered classes: normal/benign epithelium, ADH/low-grade DCIS, high-grade DCIS, and IDC.

We then trained and compared two convolutional neural network (CNN) architectures, Xception (37) and EfficientNet-B0 (38), to assess whether the observed effect was consistent across architectures rather than specific to a single one. Xception was included as a higher-capacity model for capturing subtle spatial patterns, whereas EfficientNet-B0 provided a more parameter-efficient alternative for this modest-sized cohort. Each architecture was trained separately on H&E and TRPV4 IHC image tiles using patient-level 3-fold cross-validation in the development cohort, and final performance was evaluated by applying ensembles trained on the UVA cohort to the independent GWU cohort without retraining or fine-tuning. Performance was assessed at both the tile and patient levels using macro-averaged F1-scores and areas under the receiver operating characteristic curve (ROC-AUC), together with per-class metrics and analysis of whether misclassifications occurred predominantly between adjacent diagnostic categories. This framework allowed us to test whether TRPV4 IHC, which encodes biologically relevant TRPV4 localization patterns beyond routine morphology alone, improves deep learning-based classification relative to H&E and whether any advantage generalizes to an independent external cohort.

## Materials and Methods

### Study Design and Patient Cohorts

We conducted a multi-institutional retrospective study to develop and validate deep learning models for classifying TRPV4 immunohistochemistry (IHC) and matched hematoxylin and eosin (H&E) breast pathology. The study was approved by the institutional review boards at GWU and UVA with a waiver of informed consent for de-identified archival material. Specimens were obtained in part through the National Cancer Institute’s Cooperative Human Tissue Network (CHTN; RRID: SCR_004446), coordinated at UVA.

After excluding intermediate-grade DCIS cases, the final dataset comprised 108 unique patients and 24,248 annotated tiles (299×299-pixel image patches extracted from whole-slide images). Patients were divided into a development cohort from the UVA (n=69) and an independent external test cohort from GWU (n=39) (**Fig. 1A**; **Table 1**; **Supplementary Table S1**). Because the development cohort was evaluated by patient-level 3-fold cross-validation rather than a single fixed train/validation split, we report cohort-level development and external test sizes in the main text and provide fold-level allocations in the Supplement. The GWU cohort substantially overlaps with cases previously analyzed in our mechanistic study of TRPV4 mechanotransduction in DCIS (35). Within UVA, evaluable H&E slides were available for 62 patients (9,623 tiles) and TRPV4 IHC for 51 patients (6,792 tiles). Within GWU, H&E slides were available for 31 patients (4,511 tiles) and TRPV4 IHC for 33 patients (3,322 tiles). Not all patients had both modalities available because of tissue exhaustion or staining/quality-control exclusions. Across cohorts and stain modalities, class frequencies were moderately imbalanced and differed between UVA and GWU (**Fig. 1A**; **Supplementary Table S1**).

**Figure 1.**
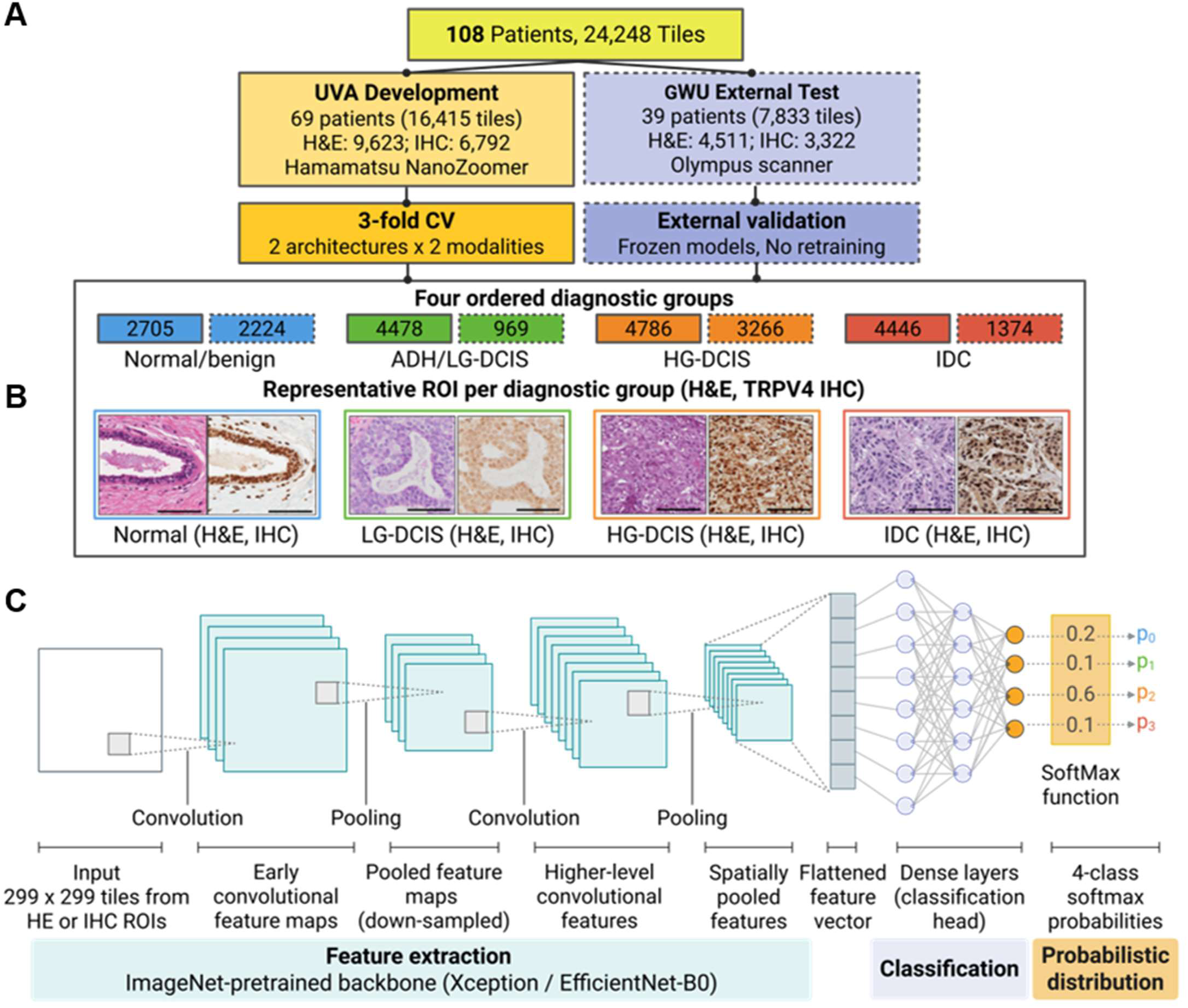
Study design and deep learning architecture. Overview of the multi-institutional dataset and deep learning pipeline for TRPV4 IHC-enhanced DCIS classification. **(A)** Patient and tile distribution across UVA development (n=69 patients) and GWU external test (n=39 patients) cohorts, stratified by stain modality (H&E, TRPV4 IHC) and diagnostic category. The four ordered diagnostic groups reflect the DCIS progression spectrum: normal/benign (blue), ADH/low-grade DCIS (green), high-grade DCIS (orange), and invasive ductal carcinoma (IDC, red). **(B)** Representative ROIs for each diagnostic group in H&E and TRPV4 IHC, illustrating typical morphology and staining patterns from which tiles were sampled. Scale bars, 200 µm. **(C)** The schematic is adapted from “Convolutional Neural Networks (CNNs)” template by Martina Maritan, created with BioRender.com. We used two ImageNet-pretrained CNN architectures (Xception and EfficientNet-B0). UVA tiles were partitioned using patient-level 3-fold cross-validation (CV) to train separate models for each stain modality and backbone architecture. Each fold comprised tiles from distinct patients, ensuring no patient appeared in multiple folds. We evaluated ensemble models (averaging predictions from the three fold-specific models per modality and architecture) on an independent external test set scanned on a different whole-slide imaging platform (Olympus VS200 at GWU vs Hamamatsu NanoZoomer at UVA), without retraining, to assess cross-platform generalizability.

**Table 1.**
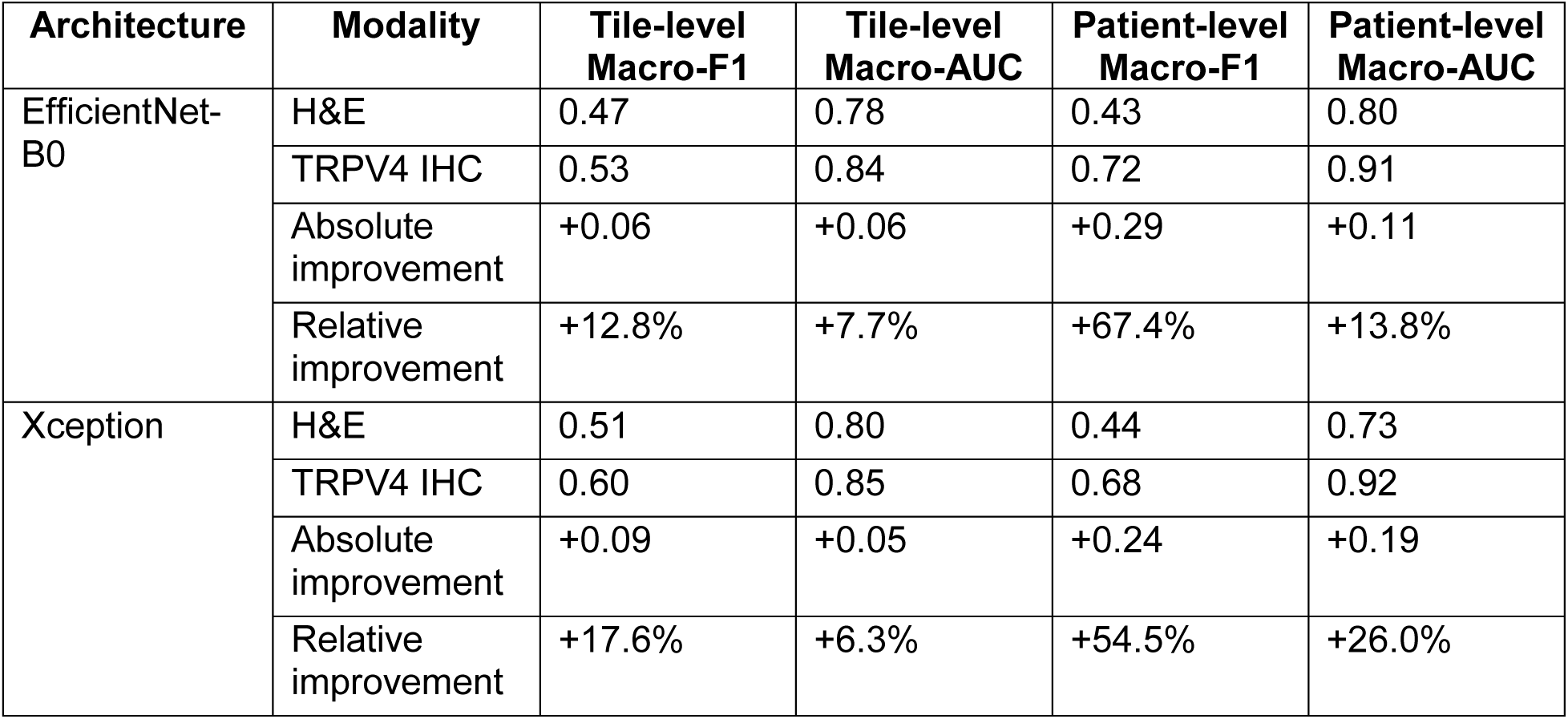
External test performance on the GWU cohort. External test performance comparing H&E and TRPV4 IHC ensemble models evaluated on the held-out GWU cohort without retraining or fine-tuning. Each ensemble comprises three fold-specific models trained on the UVA development cohort using patient-level 3-fold cross-validation, with predictions averaged across folds. Tile-level macro-F1 and macro-AUC summarize tile-level discrimination, and patient-level metrics were computed after aggregating tile-level probability outputs within each patient. Absolute and relative improvements quantify the performance gain of TRPV4 IHC over H&E for each architecture. The largest gains were observed for patient-level macro-F1, with relative improvements of 54.5% to 67.4%.

| Architecture | Modality | Tile-level Macro-F1 | Tile-level Macro-AUC | Patient-level Macro-F1 | Patient-level Macro-AUC |
| --- | --- | --- | --- | --- | --- |
| EfficientNet-B0 | H&E | 0.47 | 0.78 | 0.43 | 0.80 |
|  | TRPV4 IHC | 0.53 | 0.84 | 0.72 | 0.91 |
|  | Absolute improvement | +0.06 | +0.06 | +0.29 | +0.11 |
|  | Relative improvement | +12.8% | +7.7% | +67.4% | +13.8% |
| Xception | H&E | 0.51 | 0.80 | 0.44 | 0.73 |
|  | TRPV4 IHC | 0.60 | 0.85 | 0.68 | 0.92 |
|  | Absolute improvement | +0.09 | +0.05 | +0.24 | +0.19 |
|  | Relative improvement | +17.6% | +6.3% | +54.5% | +26.0% |

UVA cases were used exclusively for model training, hyperparameter selection, and patient-level three-fold cross-validation. GWU cases were held out entirely for external testing and were not used at any stage of training or model selection. All data partitioning was performed at the patient level to prevent information leakage. Patient IDs were assigned using stratified 3-fold cross-validation to yield folds with comparable class distributions across the four ordered diagnostic groups. This design was chosen to evaluate cross-institutional generalizability, a clinically important requirement for pathology deployment, rather than performance within a single-site proof-of-concept dataset.

### Histologic Preparation, TRPV4 IHC, and Image Acquisition

Consecutive 4-µm FFPE sections from each case were stained with routine H&E and TRPV4 IHC; the GWU cohort largely overlaps with cases from our prior mechanistic study of TRPV4 mechanotransduction (35). TRPV4 immunostaining was performed on a Ventana Discovery Ultra automated platform using anti-TRPV4 antibody (Abcam ab39260, RRID:AB_1143677; rabbit polyclonal; 1:40) with CC1 (Tris-EDTA, pH 8.0) heat-induced epitope retrieval, HRP/DAB detection (OmniMap anti-rabbit HRP; ChromoMap DAB), and hematoxylin counterstaining, as previously described (35, 36). Each run included positive-control tissue and negative controls with primary antibody omitted.

Whole-slide images were digitized at 40× magnification. UVA slides were scanned using a Hamamatsu NanoZoomer (∼0.25 µm/pixel), and GWU slides were scanned by VitroVivo Biotech (Rockville, MD) using an Olympus VS200 with comparable resolution. Three board-certified breast pathologists independently reviewed slides and annotated epithelial regions of interest (ROIs) including ducts and terminal duct-lobular units, while excluding artifacts (e.g., tissue folds). Non-overlapping 299 × 299 pixel tiles were extracted from annotated ROIs on both H&E and TRPV4 IHC slides; paired ROI-level examples are shown in **Fig. 1B**, representative paired 299 × 299 input tiles are shown in **Fig. 2**, and additional cohort-specific examples are shown in Supplementary Fig. S1.

**Figure 2.**
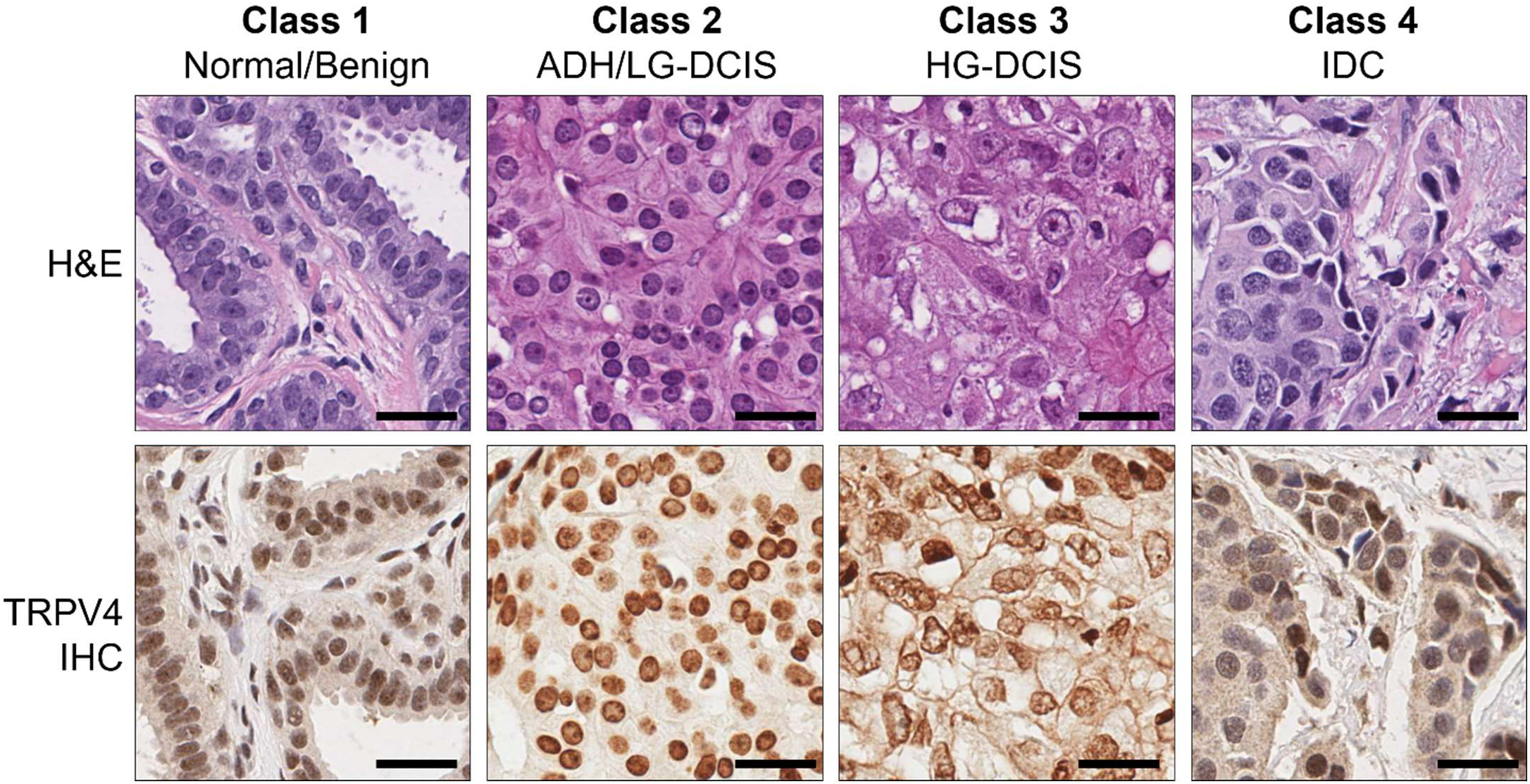
Representative 299 × 299 input tiles from the four modeled diagnostic classes. Representative paired H&E and TRPV4 IHC tiles corresponding to the four modeled diagnostic classes are shown: Class 1, normal/benign; Class 2, ADH/low-grade DCIS; Class 3, high-grade DCIS; and Class 4, invasive ductal carcinoma (IDC). In the H&E row, the examples represent tiles annotated by pathologists as normal/benign, ADH/low-grade DCIS, high-grade DCIS, and IDC, respectively. In the TRPV4 IHC row, Class 1 shows predominantly intracellular staining, Class 2 shows occasional cells with plasma membrane-associated TRPV4 staining, Class 3 shows more prevalent plasma membrane-associated staining, and Class 4 shows focal cells with plasma membrane-associated staining. Tiles were extracted from pathologist-annotated epithelial regions of interest and resized to 299 × 299 pixels for model input. Scale bars, 50 μm.

### Diagnostic Labels and Ordinal Grouping

For ROIs reviewed by multiple pathologists, the final diagnostic label was assigned by majority vote; equivocal or discordant ROIs without clear majority were excluded. Because pathologists reviewed different subsets of ROIs based on case availability, formal inter-rater reliability could not be calculated. Original labels included: normal, benign, ADH, LG-DCIS, HG-DCIS, IDC, and a small set of indeterminate ROIs annotated as intermediate-grade DCIS, which were excluded a priori. Intermediate-grade DCIS was excluded to reduce label ambiguity in this initial model-development study and to focus the primary analysis on more reproducibly defined diagnostic groups along the DCIS progression spectrum.

### Patient-Level Cross-Validation and Data Partitioning

To avoid leakage of patient-specific morphology across folds, we defined all splits at the patient-ID level. For each modality (TRPV4 IHC and H&E) within the UVA cohort, we assigned patients to 3 cross-validation folds as follows: for each patient, we computed the majority label group among that patient’s image tiles. Patients were then stratified by this majority label, and within each label group, patients were ordered by decreasing tile count and distributed in a round-robin fashion across three folds (Fold 0, 1, and 2). This yielded three patient-disjoint folds with similar class distributions at the image tile level. For each fold in turn, the held-out fold served as internal validation, and the remaining two folds were combined as the training set. All GWU patients were reserved as the external test cohort and were not included in any cross-validation procedure.

### Image Preprocessing and Data Augmentation

RGB tiles were resized to 299 × 299 pixels, and raw 8-bit RGB pixel intensities were scaled to [0,1]. During training only, we applied on-the-fly geometric and photometric augmentations to both H&E and TRPV4 IHC tiles, including random flips, 90° rotations, random zoom/cropping, color jitter, and additive Gaussian noise; augmented outputs were clipped to [0,1]. Tiles were then processed using the backbone-specific ImageNet preprocessing function. Validation and external test tiles underwent the same resizing and intensity scaling, followed by backbone-specific preprocessing, but were not augmented.

### Network Architectures and Implementation

We trained two ImageNet-pretrained CNN backbones, Xception and EfficientNet-B0, for each imaging modality, replacing the original classification heads with a shared architecture consisting of global average pooling, a 256-unit ReLU layer with L2 regularization (λ=0.001), dropout (0.6), and a 4-class output layer (**Fig. 1C; Supplementary Table S2**). Mixed-precision training (Keras mixed-float16 policy) was enabled to reduce memory use; the final layer was kept in float32 for numerical stability.

### Training Procedure, Optimization, and Class Imbalance

For each stain-backbone combination and cross-validation fold, we used a two-stage transfer-learning protocol. In Stage 1, the ImageNet-pretrained backbone was frozen and only the classification head was trained; in Stage 2, the upper backbone layers were unfrozen for fine-tuning. Models were optimized with Adam using a class-weighted, label-smoothed categorical cross-entropy objective to address class imbalance, with class weights set inversely proportional to training-set frequencies and additional upweighting of IDC to penalize missed invasive disease. Training used validation macro-F1 for learning-rate scheduling and early stopping to limit overfitting. A class-weighted focal-loss variant was explored but was not used for the primary experiments due to less stable optimization. Training was performed on an NVIDIA GeForce RTX 4090 GPU using TensorFlow 2.10.1/Keras with mixed precision (batch size=4). Depending on modality and backbone, approximate training time per model/fold ranged from about 6 to 17 minutes (**Supplementary Table S2**).

### Ensemble Inference, Tile- and Patient-Level Predictions

For each modality-architecture pair, three models (one per patient-level cross-validation fold) were trained on the UVA training cohort. For external testing, we applied the UVA-trained fold models to the GWU cohort without retraining or fine-tuning and averaged predicted class probabilities across folds to form an ensemble. Metrics were computed at both the tile and patient levels. For patient-level inference, tile probability vectors were aggregated by mean probability pooling, and the class with the highest mean probability was assigned as the patient-level prediction. No GWU data were used for training, model selection, or hyperparameter tuning. This ensemble framework was used to assess whether performance gains from TRPV4 IHC generalized across institutions rather than reflecting site-specific optimization.

### Evaluation Metrics and Statistical Analysis

Model performance was summarized using the macro-averaged F1-score (macro-F1), computed across all four classes to weight each class equally regardless of frequency. Macro-AUC (ROC-AUC) was computed from the model’s softmax probabilities using a multiclass one-vs-one framework and macro-averaged across the four classes. Tile-level macro-AUC was computed across all tiles, and patient-level macro-AUC was computed after aggregating tile probability vectors within each patient by mean-probability pooling. Sensitivity, specificity, and ROC-AUC were treated as prevalence-independent measures, whereas precision, negative predictive value, and F1-score were treated as prevalence-dependent and interpreted in conjunction with class support. For the external GWU test set, we estimated 95% confidence intervals (CI) for per-class F1 and AUC using non-parametric bootstrapping with 500 resamples. In each resample, we randomly selected patients (with replacement) and included all tiles from those patients, then recalculated both tile-level and patient-level metrics. We reported the 2.5th and 97.5th percentiles across resamples as the CI bounds.

### Error Analysis and Adjacent-Grade Misclassification

Given the ordinal progression of the four labels, we distinguished adjacent-grade errors (normal/benign vs ADH/LG-DCIS, ADH/LG-DCIS vs HG-DCIS, HG-DCIS vs IDC) from non-adjacent errors (e.g., normal/benign vs IDC). From the multiclass tile-level confusion matrix, we quantified the total number of misclassified tiles, the subset differing by exactly one step in the ordinal scale, and the fraction of all errors that were adjacent-grade, providing a clinically interpretable measure of error severity. The primary diagnostic gray zone in this study lies near the benign versus ADH/low-grade DCIS boundary, and therefore, this analysis also helped determine whether model errors remained concentrated at biologically plausible neighboring categories rather than across widely separated diagnostic groups.

## Results

### Internal Cross-Validation Performance on Development Cohort

Both Xception and EfficientNet-B0 architectures were trained separately on H&E and TRPV4 IHC tiles using patient-level 3-fold cross-validation in the UVA development cohort (**Fig. 1A-C**; **Supplementary Table S3**). Training was monitored with early stopping to reduce overfitting. Unless otherwise specified, values are reported as mean ± SD across the three validation folds. At the tile level, H&E- and TRPV4 IHC-based models showed broadly comparable macro performance (**Supplementary Table S3**). For H&E, validation tile-level macro-F1 ranged from 0.36-0.74 across folds (0.56±0.19 for EfficientNet-B0; 0.59±0.09 for Xception), with macro-AUC of 0.71-0.93 (0.84±0.11 for EfficientNet-B0; 0.85±0.07 for Xception). For TRPV4 IHC, tile-level macro-F1 ranged from 0.49-0.76 (0.58±0.08 for EfficientNet-B0; 0.68±0.07 for Xception), with macro-AUC of 0.83-0.94 (0.87±0.03 for EfficientNet-B0; 0.90±0.04 for Xception).

At the patient level, however, aggregation increased separation between modalities (**Supplementary Table S3**). H&E models achieved patient-level macro-F1 values of 0.60±0.09 for EfficientNet-B0 and 0.61±0.10 for Xception, with corresponding macro-AUC values of 0.84±0.10 and 0.89±0.07. TRPV4 IHC improved patient-level macro-F1 to 0.76±0.11 for EfficientNet-B0 and 0.75±0.09 for Xception, and increased patient-level macro-AUC to 0.97±0.02 for both architectures. Thus, although tile-level macro performance was broadly similar, TRPV4 IHC showed a clearer advantage after patient-level aggregation, suggesting that the TRPV4-associated signal becomes more stable when integrated across multiple tiles from the same case. Per-class performance on the internal validation folds likewise favored TRPV4 IHC over H&E in several diagnostically important categories (**Supplementary Table S4**). For ADH/low-grade DCIS, a clinically challenging category and the primary emphasis of this study, mean tile-level F1 increased in both architectures (EfficientNet-B0: 0.67±0.17 vs 0.65±0.16; Xception: 0.71±0.10 vs 0.62±0.12), with corresponding gains in sensitivity (EfficientNet-B0: 0.76±0.20 vs 0.70±0.10; Xception: 0.67±0.17 vs 0.61±0.01). Precision also improved for Xception (0.80±0.13 vs 0.66±0.24) and was similar for EfficientNet-B0 (0.60±0.17 vs 0.62±0.21). For high-grade DCIS, TRPV4 IHC also improved F1 in both architectures (EfficientNet-B0: 0.56±0.16 vs 0.51±0.33; Xception: 0.69±0.08 vs 0.58±0.19), together with higher sensitivity and precision. For invasive ductal carcinoma, mean tile-level F1 was unchanged for EfficientNet-B0 (0.50±0.27 vs 0.50±0.27) but increased for Xception (0.67±0.29 vs 0.57±0.24), with precision increasing in both architectures (EfficientNet-B0: 0.94±0.09 vs 0.86±0.13; Xception: 0.93±0.09 vs 0.76±0.07). Overall, the internal cross-validation results suggested that TRPV4 IHC provided the greatest benefit after patient-level aggregation and in diagnostically challenging categories, particularly ADH/low-grade DCIS.

### External Test Performance: TRPV4 IHC versus H&E

To assess cross-institutional generalizability, we evaluated ensembles formed by averaging predictions from the three UVA cross-validation fold models for each architecture and modality on the independent GWU cohort, without retraining or fine-tuning (**Fig. 1A,B**; **Table 1**; **Fig. 3A,B**). At the tile level (**Fig. 3A**; **Table 1**), TRPV4 IHC improved macro-level performance relative to H&E across both architectures. H&E ensembles achieved macro-F1 values of 0.47 for EfficientNet-B0 and 0.51 for Xception, with corresponding macro-AUC values of 0.78 and 0.80. TRPV4 IHC improved tile-level macro-F1 to 0.53 for EfficientNet-B0 and 0.60 for Xception, with macro-AUC increasing to 0.84 and 0.85, respectively (**Table 1**).

**Figure 3.**
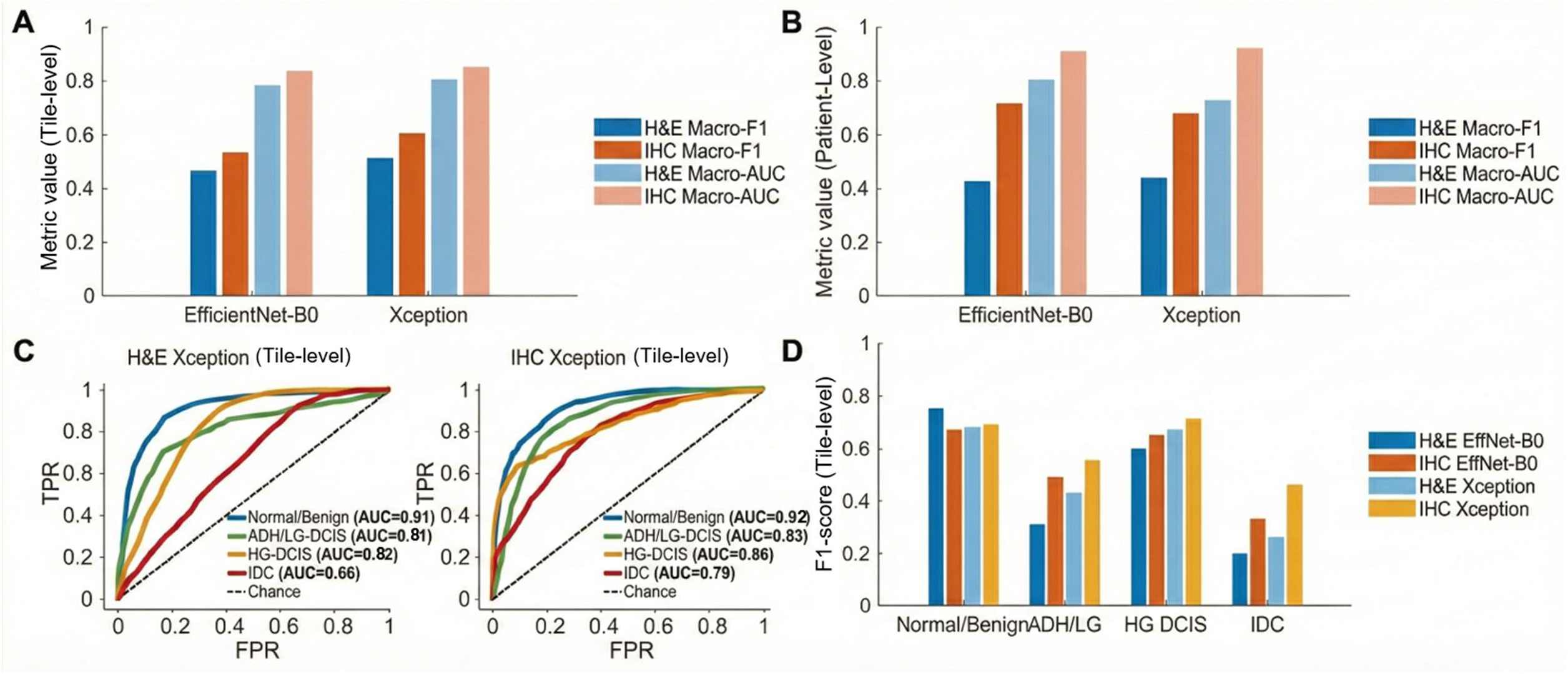
External test performance on the GWU cohort. The GWU external test cohort comprised 39 unique patients; H&E was available for 31 patients (4,511 tiles) and TRPV4 IHC for 33 patients (3,322 tiles), totaling 7,833 tiles after excluding intermediate-grade cases. **(A)** Tile-level macro-F1 and macro-AUC for H&E and TRPV4 IHC ensemble models across EfficientNet-B0 and Xception architectures (**Table 1**). **(B)** Patient-level macro-F1 and macro-AUC after aggregating tile probabilities within each patient, showing larger performance gains for TRPV4 IHC than for H&E (**Table 1**). **(C)** Tile-level one-vs-rest ROC curves for the Xception ensembles (left: H&E; right: TRPV4 IHC), illustrating improved discrimination for HG-DCIS and IDC and a modest gain for ADH/low-grade DCIS (**Table 2**). Xception is shown because it achieved marginally higher tile-level macro-F1 than EfficientNet-B0 on GWU (**Table 1**). **(D)** Tile-level per-class F1 scores for all model–modality combinations, highlighting the largest improvements for ADH/low-grade DCIS and IDC (**Table 2**).

**Table 2.** Tile-level class-wise performance on the GWU external test cohort. Per-class sensitivity, specificity, precision, F1-score, and one-vs-rest AUC are shown for ensemble models trained on the UVA cohort and evaluated on GWU without retraining. F1-score and AUC are reported with 95% confidence intervals estimated by bootstrap resampling (n=500). Sensitivity, specificity, and AUC are prevalence-independent measures, whereas precision and F1-score are prevalence-dependent; the number of evaluated tiles per class is therefore shown to indicate class support for each modality.

| Model | Class | Sensitivity | Specificity | Precision | F1-score<br>[95% CI] | AUC<br>[95% CI] | n<br>tiles |
| --- | --- | --- | --- | --- | --- | --- | --- |
| **H&E**<br>**Efficient**<br>**Net-B0** | Normal/<br>benign | 0.88 | 0.78 | 0.66 | 0.75<br>[0.74–<br>0.77] | 0.92<br>[0.91–<br>0.93] | 1,45<br>4 |
|  | ADH/LG<br>-DCIS | 0.48 | 0.81 | 0.23 | 0.31<br>[0.28–<br>0.34] | 0.70<br>[0.68–<br>0.74] | 474 |
|  | HG-DCIS | 0.50 | 0.89 | 0.75 | 0.60<br>[0.58–<br>0.62] | 0.85<br>[0.84–<br>0.86] | 1,78<br>6 |
|  | IDC | 0.15 | 0.93 | 0.31 | 0.20<br>[0.17–<br>0.23] | 0.65<br>[0.63–<br>0.67] | 797 |
| **IHC**<br>**Efficient**<br>**Net-B0** | Normal/<br>benign | 0.75 | 0.86 | 0.61 | 0.67<br>[0.65–<br>0.70] | 0.91<br>[0.90–<br>0.92] | 770 |
|  | ADH/LG<br>-DCIS | 0.73 | 0.78 | 0.36 | **0.49**<br>[0.45–<br>0.52] | **0.84**<br>[0.82–<br>0.86] | 495 |
|  | HG-DCIS | 0.55 | 0.87 | 0.77 | **0.65**<br>[0.62–<br>0.67] | 0.85<br>[0.84–<br>0.87] | 1,48<br>0 |
|  | IDC | 0.25 | 0.94 | 0.46 | **0.33**<br>[0.29–<br>0.37] | **0.74**<br>[0.72–<br>0.76] | 577 |
| **H&E**<br>**Xception** | Normal/<br>benign | 0.58 | 0.94 | 0.83 | 0.68<br>[0.66–<br>0.71] | 0.91<br>[0.90–<br>0.92] | 1,45<br>4 |
|  | ADH/LG<br>-DCIS | 0.58 | 0.88 | 0.35 | 0.43<br>[0.40–<br>0.47] | 0.81<br>[0.79–<br>0.84] | 474 |
|  | HG-DCIS | 0.69 | 0.76 | 0.65 | 0.67<br>[0.65–<br>0.69] | 0.82<br>[0.81–<br>0.84] | 1,78<br>6 |
|  | IDC | 0.27 | 0.84 | 0.26 | 0.26<br>[0.24–<br>0.29] | 0.66<br>[0.65–<br>0.68] | 797 |
| **IHC**<br>**Xception** | Normal/<br>benign | 0.67 | 0.92 | 0.71 | **0.69**<br>[0.66–<br>0.72] | **0.92**<br>[0.91–<br>0.93] | 770 |
|  | ADH/LG<br>-DCIS | 0.65 | 0.88 | 0.49 | **0.56**<br>[0.52–<br>0.59] | **0.83**<br>[0.81–<br>0.86] | 495 |
|  | HG-DCIS | 0.65 | 0.86 | 0.79 | **0.71**<br>[0.69–<br>0.73] | **0.86**<br>[0.85–<br>0.88] | 1,48<br>0 |
|  | IDC | 0.52 | 0.84 | 0.41 | **0.46**<br>[0.42–<br>0.49] | **0.79**<br>[0.76–<br>0.80] | 577 |

The largest gains emerged at the patient level (**Fig. 3B**; **Table 1**), which is the more clinically relevant level of interpretation. H&E ensembles achieved patient-level macro-F1 values of 0.43 for EfficientNet-B0 and 0.44 for Xception, with macro-AUC values of 0.80 and 0.73, respectively. In contrast, TRPV4 IHC improved patient-level macro-F1 to 0.72 for EfficientNet-B0 and 0.68 for Xception, with macro-AUC increasing to 0.91 and 0.92. Thus, the advantage of TRPV4 IHC became more pronounced after patient-level aggregation, consistent with a workflow in which diagnoses are made by integrating information across multiple regions rather than individual tiles. Class-wise tile-level discrimination is shown by ROC curves in **Fig. 3C**. In the Xception comparison shown, TRPV4 IHC improved per-class AUC relative to H&E for high-grade DCIS (0.86 vs 0.82) and IDC (0.79 vs 0.66), while macro-AUC increased from 0.80 to 0.85 (**Table 1**; **Table 2**). At the tile level, the gain for ADH/low-grade DCIS was more modest in this architecture (0.83 vs 0.81) (**Table 2**), suggesting that the principal benefit in this diagnostically challenging category becomes clearer after broader patient-level integration.

Tile-level per-class F1-scores similarly showed consistent TRPV4 IHC gains across architectures (**Fig. 3D**; **Table 2**), with the largest increases for ADH/low-grade DCIS (0.31-0.43 to 0.49-0.56) and IDC (0.20-0.26 to 0.33-0.46). Notably, patient-level per-class analyses further emphasized this pattern, with the largest AUC gains seen for ADH/low-grade DCIS and IDC on the GWU cohort (**Fig. 3C**; **Supplementary Table S5**). Together, these results indicate that TRPV4 IHC improves external-test discrimination overall, with the clearest clinical benefit in diagnostically challenging categories.

### Patient-level per-class performance comparison

**Fig. 4** summarizes patient-level sensitivity (**Fig. 4A**), precision (**Fig. 4B**), and AUC (**Fig. 4C**) by diagnostic category for all four modality-architecture combinations on the GWU external test cohort (**Supplementary Table S5**). Because patient-level classification more closely reflects diagnostic practice than tile-level predictions, these analyses provide the most clinically relevant view of model behavior across lesion categories.

**Figure 4.**
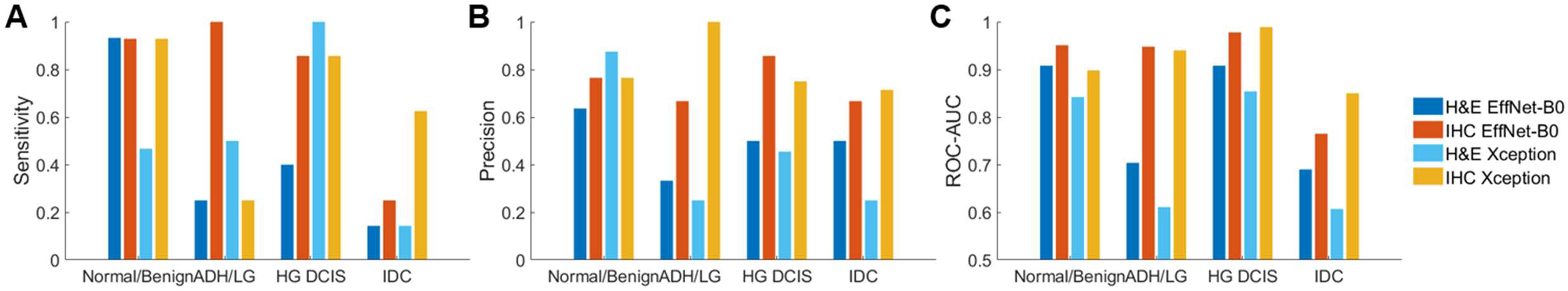
Patient-level per-class performance comparison on the GWU external test cohort. Patient-level sensitivity **(A)**, precision **(B)**, and one-vs-rest ROC-AUC **(C)** bpay diagnostic category for all four model–modality combinations (H&E EfficientNet-B0, TRPV4 IHC EfficientNet-B0, H&E Xception, IHC Xception). The GWU cohort comprised 39 patients (H&E n=31; TRPV4 IHC n=33). IHC showed higher AUC than H&E across categories, with the largest gains for ADH/low-grade DCIS (IHC 0.94–0.95 vs H&E 0.61–0.70) and IDC (IHC 0.77–0.85 vs H&E 0.61–0.69). Exact values and 95% confidence intervals are provided in **Supplementary Table S5**.

Sensitivity (**Fig. 4A**) showed architecture-dependent patterns, whereas AUC (**Fig. 4C**) consistently favored TRPV4 IHC over H&E, with the largest gains for ADH/low-grade DCIS (IHC 0.94-0.95 vs H&E 0.61–0.70) and IDC (IHC 0.77-0.85 vs H&E 0.61–0.69) (**Supplementary Table S5**). These two categories are clinically important because they include a diagnostically difficult low-grade boundary and the transition to overt invasion.

Sensitivity improvements were most prominent for EfficientNet-B0 in ADH/low-grade DCIS (IHC 1.00 vs H&E 0.25) and high-grade DCIS (IHC 0.86 vs H&E 0.40), while Xception showed the largest sensitivity gain for IDC (IHC 0.63 vs H&E 0.14) (**Supplementary Table S5**).

Precision (**Fig. 4B**) was also higher for IHC than H&E in ADH/low-grade DCIS (IHC 0.67-1.00 vs H&E 0.25-0.33), high-grade DCIS (IHC 0.75-0.86 vs H&E 0.46-0.50), and IDC (IHC 0.67-0.71 vs H&E 0.25-0.50), although precision is prevalence-dependent and should therefore be interpreted alongside sensitivity, AUC, and class support (**Supplementary Table S5**). H&E Xception achieved the highest precision for normal/benign cases (0.88), reflecting a trade-off between specificity for normal tissue and sensitivity for abnormal classes.

### Error Pattern Analysis

To characterize model failure modes, we examined tile-level confusion matrices for all four modality-architecture combinations on the external GWU test cohort (**Supplementary Fig. S2**). Across all models, most errors reflected confusion between adjacent diagnostic categories along the DCIS progression axis rather than between widely separated classes. Adjacent-grade errors accounted for 68.3% (1359/1991) and 79.0% (1538/1947) of misclassified tiles for the H&E models (**Supplementary Fig. S2A,C**), and 72.8% (1031/1417) and 71.0% (875/1232) for the TRPV4 IHC models (**Supplementary Fig. S2B,D**). Overall, model errors predominantly occurred between neighboring diagnostic groups, consistent with biologically plausible decision boundaries along a graded DCIS progression spectrum. This pattern suggests that when the models were incorrect, they were usually wrong by one diagnostic step rather than by implausibly large category jumps.

## Discussion

Using patient-level cross-validation on an internal development cohort and a fully held-out external test cohort from a different institution and scanner, we found that CNNs trained on TRPV4 IHC tiles outperformed H&E-based models across both architectures, with the largest gains emerging at the patient level. On the external GWU cohort, patient-level macro-F1 improved from 0.43-0.44 with H&E to 0.68-0.72 with TRPV4 IHC, and patient-level macro-AUC improved from 0.73-0.80 to 0.91-0.92. These patient-level gains are particularly relevant for pathology practice, where diagnoses are made by integrating information across multiple regions rather than isolated image tiles. Per-class analyses further showed that the largest discrimination gains were observed for ADH/low-grade DCIS and IDC, two clinically consequential categories that are often challenging to distinguish at the microscope. For ADH/low-grade DCIS in particular, tile-level gains were modest in some analyses but became more pronounced after patient-level aggregation, suggesting that the TRPV4-associated signal becomes more reliable when integrated across multiple ROIs from the same case.

These findings extend prior DCIS-related deep learning work, which has largely relied on morphologic features from H&E to distinguish benign from atypical or malignant lesions or to separate DCIS from invasive disease (23–31). Such approaches can be informative, but they necessarily inherit the ambiguity of morphology-based grading, particularly at clinically important boundaries. Here, we instead evaluate a mechanistically motivated, pathology-readable stain linked to crowding-induced mechanotransduction in confined ducts. In our prior mechanistic work (35), plasma membrane-associated TRPV4 was linked to a pro-invasive mechanotransduction program, and in related analytical-validation work from our group (36), pathologists were able to score plasma membrane-associated TRPV4 with high interobserver reproducibility (weighted Fleiss’ κ=0.823), supporting the idea that this signal is interpretable by humans as well as by algorithms. Taken together, these observations support TRPV4 IHC not as a replacement for pathologist judgment, but as a biologically grounded complement to morphology that may help improve classification in diagnostically challenging cases. To place the present findings in context, selected prior deep learning studies (23–25, 29–31) directly relevant to DCIS-related breast histopathology are summarized in **Table 3**.

**Table 3.** Comparison of the present study with selected deep learning studies in DCIS-related breast histopathology. Studies were selected for direct relevance to DCIS-related breast histopathology, including lesion classification, grading, biopsy upstaging prediction, and invasive recurrence prediction. Direct numerical comparison should be interpreted cautiously because tasks, labels, endpoints, and validation designs differ across studies.

| Study | Task | Dataset | Main result |
| --- | --- | --- | --- |
| **This study** | 4-class classification: normal/benign, ADH/LG-DCIS, HG-DCIS, IDC | 108 patients total (69 in development, 39 in external validation); 24,248 annotated tiles | TRPV4 IHC: macro-AUC 0.91–0.92; macro-F1 0.68–0.72. H&E: macro-AUC 0.73–0.80; macro-F1 0.43–0.44 |
| **Doyle et al., 2025** | Prediction of invasive recurrence after pure DCIS | 652 patients total (558 in development, 94 in external validation) | AUC 0.75 |
| **Huang et al., 2024** | Prediction of biopsy upstaging in ADH/DCIS | 733 patients total (592 in development, 141 in validation) | Deep learning alone, AUC 0.81; with clinical variables, AUC 0.87 |
| **Kanavati et al., 2022** | 3-class classification: benign, DCIS, IDC | 3,672 whole-slide images from 2 institutions | AUC 0.960 for DCIS; AUC 0.977 for IDC |
| **Wetstein et al., 2021** | DCIS nuclear grading, grades 1, 2, and 3 | 109 patients total (59 in development, 50 in independent test set) | Agreement with expert pathologists; kappa 0.70–0.77 |
| **Araujo et al., 2017** | 4-class classification: normal, benign, in situ, invasive | 285 images from a single institution | 77.8% accuracy for 4-class classification |
| **Bejnordi et al., 2016** | Binary detection of DCIS versus benign tissue | 205 whole-slide images from 2 datasets | FROC reported; AUC not reported |

This study was intentionally designed to emphasize generalizability rather than engineering optimization. The performance gains observed with TRPV4 IHC were maintained when models trained on the UVA cohort were applied without retraining or fine-tuning to a fully independent GWU cohort scanned on a different platform. This cross-institutional robustness is clinically more important than scanner-specific optimization, because any pathology decision-support tool must perform across multiple laboratories and imaging workflows to have realistic translational value. The fact that the TRPV4 advantage was observed in both Xception and EfficientNet-B0 further suggests that the signal is not specific to one single architecture, but instead reflects a transferable biological feature.

The present findings also help clarify why TRPV4 IHC may be useful in a tile-based framework. Tile-level analysis inevitably loses some architectural context that pathologists use in routine diagnosis, and this is an important limitation of the present study. At the same time, localized TRPV4 staining patterns may partially compensate for this loss of context by encoding biologically meaningful lesional information at the cellular and ductal level. Consistent with this interpretation, most model errors remained concentrated between adjacent diagnostic categories along the DCIS progression axis rather than between widely separated classes, supporting biologically plausible decision boundaries. From a pathology-facing perspective, the most realistic role for this approach is as a decision-support tool rather than a stand-alone classifier. A TRPV4 IHC-assisted model could help identify foci or cases that warrant closer review, particularly in diagnostically ambiguous low-grade lesions or when concern for occult invasion is heightened. While sensitivity and precision varied by architecture, the consistent AUC improvements suggest that TRPV4 IHC encodes a robust, transferable signal that improves discrimination across architectures.

This study has several limitations. The number of patients, particularly in the external cohort for ADH/LG-DCIS, was modest, leading to wide confidence intervals and limiting the precision of per-class estimates (**Supplementary Table S5**). Although we evaluated generalization across institutions and scanners, the external validation was performed at a single independent site, and additional multi-site testing will strengthen confidence in broader deployment. Intermediate-grade DCIS and morphologic mimics were excluded to reduce label noise, but remain clinically important and should be incorporated in future work. Finally, this study was not designed to assess prognostic value or treatment impact; prospective validation with outcome-linked endpoints will be essential before clinical translation.

Future work should include prospective validation, expansion to larger multi-institutional cohorts, and transition from tile-based classification to whole-slide inference. It will also be important to evaluate how TRPV4 IHC performs alongside other clinicopathologic variables and biomarkers rather than in isolation. Because the greatest gains in the current study were observed in diagnostically challenging categories, future studies should specifically test whether TRPV4-assisted models can improve reproducibility and diagnostic confidence at the benign versus ADH/low-grade DCIS boundary. Despite these limitations, our findings provide proof-of-concept that TRPV4 IHC paired with deep learning can enhance DCIS-focused classification in a multi-institutional setting. The largest gains in discrimination were observed in ADH/LG-DCIS and IDC, which align with clinically difficult transitions and support the value of biologically grounded stains in diagnostic gray zones. More broadly, these results suggest that mechanosensitive biomarkers such as TRPV4 may help bridge cellular mechanics and AI-assisted pathology, offering a path toward more interpretable and biologically informed diagnostic models.

## Funding

This work was supported by the Elsa Pardee Foundation Award, the George Washington University Technology Maturation Award, the George Washington Cancer Center, and the Katzen Cancer Research Pilot Award. The funders had no role in study design; data collection, analysis, or interpretation; or manuscript preparation.

## Data availability

De-identified data and analysis code will be provided by the corresponding author upon reasonable request, subject to institutional policies and data use agreements.

## Authors’ contributions

I. J. Yoo: implemented the initial Xception model; wrote the initial prototype code to train the model on H&E versus IHC images; curated the image data. R. Karthikeyan: used Xception as a backbone and implemented EfficientNet-B0 in the pipeline; curated the image data. K. Kamat: data curation; coded the image input process; image summarization. C. Chan: data curation; image summarization. S. Samankan: ROI pathology annotation. E. Arbzadeh: ROI pathology annotation. A. Schwartz: ROI pathology annotation. P. Latham: identification of additional GWU cases. I. Chung: conceptualization; study design; methodology; final model development and optimization; formal analysis; supervision; project administration; funding acquisition; writing (original draft); writing (review and editing). All authors read and approved the final manuscript.

## Acknowledgements

Tissue samples were provided by the Cooperative Human Tissue Network (CHTN), which is funded by the National Cancer Institute; other investigators may have received specimens from the same subjects. We thank Rebecca Blackwell (CHTN, University of Virginia) for case identification; Dr. Christopher Moskaluk (CHTN, University of Virginia) for histologic review; Dr. Patcharin Pramoonjago and Barushi Amarasinghe (Biorepository and Tissue Research Facility, University of Virginia) and Dr. Johnson Wang (VitroVivo Biotech, Inc, Rockville, MD) for tissue staining; and Ken Schill (DigitalScope) for assistance with IHC image data management.

## Conflict of interest

I. Chung is an inventor on U.S. Patent No. 12,013,398 related to plasma-membrane relocalization based diagnostics. The patent is assigned to The George Washington University, is not licensed, and no commercial products are currently in development. No authors have received or will receive financial compensation related to this patent. The authors declare no other competing interests.

## Notes

### Author Declarations

Institutional Review Board of The George Washington University gave ethical approval for this work and waived informed consent for the use of de-identified archival breast pathology material (protocol NCR203065, Targeting biomechanical alterations in ductal carcinoma in situ). Only authorized study personnel had access to direct identifiers for linkage purposes. Identifiers were replaced with study codes prior to analysis, and all model development and evaluation were performed on coded/de-identified data. Institutional Review Board of the University of Virginia gave ethical approval for this work and waived informed consent for the use of de-identified archival breast pathology material. Samples were provided by the National Cancer Institute Cooperative Human Tissue Network and were received by the study team in coded/de-identified form.

### Summary of Updates

This revised version improves the manuscript's clarity, pathology framing, and clinical focus. The revision clarifies the primary emphasis on the benign versus ADH/low-grade DCIS boundary, strengthens the pathology-facing presentation of representative H&E and TRPV4 IHC examples, adds a summary table of deep learning architecture and training settings, clarifies dataset composition and patient-level cross-validation, adds a comparison table with related DCIS-relevant deep learning studies, and substantially revises the Discussion, limitations, and future-work sections.

