## Supplementary Tables and Figures for "Mechanosensitive TRPV4 immunohistochemistry improves deep learning-based classification of ductal carcinoma in situ beyond H&E morphology"

| Characteristic | UVA (Development) | GWU (External Test) | Total |
|---|---|---|---|
| **Unique patients** | 69 | 39 | **108** |
| Patients with H&E | 62 | 31 | 93 |
| Patients with TRPV4 IHC | 51 | 33 | 84 |
| Patients with both modalities | 44 | 25 | 69 |
| **Total tiles (after intermediate-grade DCIS exclusion)** | 16,415 | 7,833 | **24,248** |
| **H&E tiles** | **9,623 (58.6%)** | **4,511 (57.6%)** | **14,134 (58.3%)** |
| Normal/benign | 1,367 (14.2%) | 1,454 (32.2%) | 2,821 (20.0%) |
| ADH/low-grade DCIS | 2,878 (29.9%) | 474 (10.5%) | 3,352 (23.7%) |
| High-grade DCIS | 3,252 (33.8%) | 1,786 (39.6%) | 5,038 (35.6%) |
| IDC | 2,126 (22.1%) | 797 (17.7%) | 2,923 (20.7%) |
| **TRPV4 IHC tiles** | **6,792 (41.4%)** | **3,322 (42.4%)** | **10,114 (41.7%)** |
| Normal/benign | 1,338 (19.7%) | 770 (23.2%) | 2,108 (20.8%) |
| ADH/low-grade DCIS | 1,600 (23.6%) | 495 (14.9%) | 2,095 (20.7%) |
| High-grade DCIS | 1,534 (22.6%) | 1,480 (44.5%) | 3,014 (29.8%) |
| IDC | 2,320 (34.2%) | 577 (17.4%) | 2,897 (28.6%) |
| **Cross-validation folds (UVA only)** | | | |
| H&E: Fold sizes (patients) | 22, 21, 19 | — | — |
| IHC: Fold sizes (patients) | 19, 16, 16 | — | — |

**Supplementary Table S1. Cohort characteristics and tile-level dataset composition.**

Patient- and tile-level characteristics for the UVA development cohort and GWU external test cohort after excluding intermediate-grade DCIS cases. Overall, 108 unique patients contributed 24,248 tiles across H&E and TRPV4 IHC. Modality availability varied by patient due to tissue exhaustion and/or quality-control exclusions; therefore, per-modality patient counts and fold sizes differ (UVA). The UVA cohort was used for patient-level three-fold cross-validation, and the GWU cohort served as a fully held-out external test set for all models. Class distributions reflect real-world diagnostic case mix, with normal/benign and ADH/low-grade DCIS under-represented relative to high-grade DCIS. Percentages in parentheses denote the proportion within each institution and modality.

| Parameter | Setting |
|---|---|
| Task | 4-class classification (normal/benign, ADH/LG-DCIS, HG-DCIS, IDC) |
| Modalities | H&E and TRPV4 IHC |
| Development cohort | UVA; patient-level 3-fold cross-validation |
| External test cohort | GWU; fully held out |
| Input | Non-overlapping ROI-derived image tiles |
| Tile size | 299 × 299 pixels |
| Backbones | Xception; EfficientNet-B0 |
| Initialization | ImageNet pretrained |
| Classification head | Global average pooling, Dense (256, ReLU, L2 = 0.001), Dropout (0.6), and then 4-class softmax |
| Transfer learning | Two-stage training: frozen backbone, then fine-tuning |
| Optimizer | Adam |
| Loss | Class-weighted, label-smoothed categorical cross-entropy |
| Class imbalance handling | Inverse-frequency class weighting with additional IDC upweighting |
| Alternative loss explored | Class-weighted focal loss (not used for primary analyses) |
| Augmentation | Random flips, 90° rotations, zoom/cropping, color jitter, Gaussian noise |
| Precision policy | Mixed precision (mixed-float16); final layer in float32 |
| Batch size | 4 |
| Software | TensorFlow 2.10.1 / Keras |
| Hardware | NVIDIA GeForce RTX 4090 GPU |
| Approximate training time | 6-17 min per model per fold |
| Ensemble inference | Mean probability across 3 fold-specific models |
| Patient-level prediction | Mean probability pooling across tiles |
| Evaluation levels | Tile level and patient level |
| Main summary metrics | Macro-F1 and macro-AUC |
| Uncertainty estimation | Non-parametric bootstrap, 500 resamples |
| Error analysis | Adjacent-grade vs non-adjacent misclassification |

**Supplementary Table S2. Summary of deep learning architecture, training, and inference settings**

Principal deep learning settings used for H&E and TRPV4 IHC classification. Models were trained on the UVA development cohort using patient-level 3-fold cross-validation and evaluated on the fully held-out GWU external test cohort without retraining or fine-tuning.

| Architecture | Modality | Fold | Tile-level Macro-F1 | Tile-level Macro-AUC | Patient-level Macro-F1 | Patient-level Macro-AUC | n patients |
|---|---|---|---|---|---|---|---|
| **EfficientNet-B0** | H&E | 0 | 0.59 | 0.89 | 0.60 | 0.89 | 22 |
| | | 1 | 0.36 | 0.71 | 0.51 | 0.73 | 21 |
| | | **2** | **0.74** | **0.92** | **0.69** | **0.90** | 19 |
| | | Mean± SD | 0.56± 0.19 | 0.84± 0.11 | 0.60± 0.09 | 0.84± 0.10 | — |
| | **TRPV4 IHC** | 0 | 0.49 | 0.83 | 0.64 | 0.95 | 19 |
| | | **1** | **0.65** | **0.90** | **0.84** | **0.98** | 16 |
| | | 2 | 0.60 | 0.88 | 0.79 | 0.98 | 16 |
| | | Mean± SD | 0.58± 0.08 | 0.87± 0.03 | 0.76± 0.11 | 0.97± 0.02 | — |
| **Xception** | H&E | 0 | 0.57 | 0.83 | 0.64 | 0.93 | 22 |
| | | 1 | 0.51 | 0.80 | 0.49 | 0.81 | 21 |
| | | **2** | **0.69** | **0.93** | **0.69** | **0.93** | 19 |
| | | Mean± SD | 0.59± 0.09 | 0.85± 0.07 | 0.61± 0.10 | 0.89± 0.07 | — |
| | **TRPV4 IHC** | **0** | **0.64** | **0.90** | **0.84** | **0.98** | 19 |
| | | 1 | 0.76 | 0.94 | 0.66 | 0.95 | 16 |
| | | 2 | 0.64 | 0.87 | 0.74 | 0.97 | 16 |
| | | Mean± SD | 0.68± 0.07 | 0.90± 0.04 | 0.75± 0.09 | 0.97± 0.02 | — |

**Supplementary Table S3. Internal cross-validation performance (UVA cohort).**

Detailed three-fold, patient-stratified cross-validation results on the UVA development cohort. Each row reports performance on the held-out validation fold. For each architecture–modality combination, the fold with the best patient-level performance is highlighted in bold (based on patient-level macro-F1, with patient-level macro-AUC as a tie-breaker when needed). For external testing, predictions are generated by ensembling the three fold-specific models per modality and architecture (see **Table 1**). Fold sizes vary due to patient-level stratification. SD = standard deviation across three folds.

**H&E-EfficientNet-B0**

| Class | Sensitivity (mean ± SD) | Precision (mean ± SD) | F1-score (mean ± SD) | VAL tiles (fold0/fold1/fold2) |
|---|---|---|---|---|
| **Normal/Benign** | 0.90 ± 0.06 | 0.44 ± 0.13 | 0.59 ± 0.12 | 591/397/379 |
| **ADH/LG-DCIS** | 0.70 ± 0.10 | 0.62 ± 0.21 | 0.65 ± 0.16 | 1144/984/750 |
| **HG-DCIS** | 0.45 ± 0.34 | 0.59 ± 0.30 | 0.51 ± 0.33 | 1560/856/836 |
| **IDC** | 0.40 ± 0.25 | 0.86 ± 0.13 | 0.50 ± 0.27 | 1012/731/383 |

**TRPV4 IHC-EfficientNet-B0**

| Class | Sensitivity (mean ± SD) | Precision (mean ± SD) | F1-score (mean ± SD) | VAL tiles (fold0/fold1/fold2) |
|---|---|---|---|---|
| **Normal/Benign** | 0.84 ± 0.11 | 0.48 ± 0.10 | 0.60 ± 0.07 | 734/322/282 |
| **ADH/LG-DCIS** | 0.76 ± 0.20 | 0.60 ± 0.17 | 0.67 ± 0.17 | 645/501/454 |
| **HG-DCIS** | 0.51 ± 0.12 | 0.65 ± 0.26 | 0.56 ± 0.16 | 658/503/373 |
| **IDC** | 0.37 ± 0.28 | 0.94 ± 0.09 | 0.50 ± 0.27 | 1184/686/450 |

**H&E-Xception**

| Class | Sensitivity (mean ± SD) | Precision (mean ± SD) | F1-score (mean ± SD) | VAL tiles (fold0/fold1/fold2) |
|---|---|---|---|---|
| **Normal/Benign** | 0.81 ± 0.05 | 0.48 ± 0.12 | 0.59 ± 0.09 | 591/397/379 |
| **ADH/LG-DCIS** | 0.61 ± 0.01 | 0.66 ± 0.24 | 0.62 ± 0.12 | 1144/984/750 |
| **HG-DCIS** | 0.58 ± 0.29 | 0.63 ± 0.08 | 0.58 ± 0.19 | 1560/856/836 |
| **IDC** | 0.49 ± 0.28 | 0.76 ± 0.07 | 0.57 ± 0.24 | 1012/731/383 |

**TRPV4 IHC-Xception**

| Class | Sensitivity (mean ± SD) | Precision (mean ± SD) | F1-score (mean ± SD) | VAL tiles (fold0/fold1/fold2) |
|---|---|---|---|---|
| **Normal/Benign** | 0.91 ± 0.07 | 0.52 ± 0.12 | 0.65 ± 0.09 | 734/322/282 |
| **ADH/LG-DCIS** | 0.67 ± 0.17 | 0.80 ± 0.13 | 0.71 ± 0.10 | 645/501/454 |
| **HG-DCIS** | 0.69 ± 0.18 | 0.74 ± 0.19 | 0.69 ± 0.08 | 658/503/373 |
| **IDC** | 0.57 ± 0.37 | 0.93 ± 0.09 | 0.67 ± 0.29 | 1184/686/450 |

**Supplementary Table S4. Per-class internal cross-validation performance (UVA cohort).**

Per-class sensitivity, precision, and F1 score are computed on validation tiles for each held-out fold of patient-stratified 3-fold cross-validation. Values are reported as mean ± SD across three folds. The final column lists the number of validation tiles per fold (fold0/fold1/fold2).

| Model | Class | Sensitivity | Specificity | Precision | F1 [95% CI] | AUC [95% CI] | n patients |
|---|---|---|---|---|---|---|---|
| H&E EfficientNet-B0 | Normal/ benign | 0.93 | 0.50 | 0.64 | 0.76 [0.56–0.89] | 0.91 [0.80–0.99] | 15 |
| | ADH/LG-DCIS | 0.25 | 0.93 | 0.33 | 0.29 [0.00–0.67] | 0.70 [0.41–0.97] | 4 |
| | HG-DCIS | 0.40 | 0.92 | 0.50 | 0.44 [0.00–0.83] | 0.91 [0.77–1.00] | 5 |
| | IDC | 0.14 | 0.96 | 0.50 | 0.22 [0.00–0.60] | 0.69 [0.48–0.87] | 7 |
| IHC EfficientNet-B0 | Normal/ benign | 0.93 | 0.79 | 0.77 | 0.84 [0.67–0.97] | 0.95 [0.85–1.00] | 14 |
| | ADH/LG-DCIS | 1.00 | 0.93 | 0.67 | 0.80 [0.33–1.00] | 0.95 [0.82–1.00] | 4 |
| | HG-DCIS | 0.86 | 0.96 | 0.86 | 0.86 [0.55–1.00] | 0.98 [0.92–1.00] | 7 |
| | IDC | 0.25 | 0.96 | 0.67 | 0.36 [0.00–0.71] | 0.77 [0.58–0.96] | 8 |
| H&E Xception | Normal/ benign | 0.47 | 0.94 | 0.88 | 0.61 [0.35–0.82] | 0.84 [0.68–0.97] | 15 |
| | ADH/LG-DCIS | 0.50 | 0.78 | 0.25 | 0.33 [0.00–0.67] | 0.61 [0.24–1.00] | 4 |
| | HG-DCIS | 1.00 | 0.77 | 0.46 | 0.63 [0.31–0.86] | 0.85 [0.70–0.98] | 5 |
| | IDC | 0.14 | 0.88 | 0.25 | 0.18 [0.00–0.50] | 0.61 [0.38–0.82] | 7 |
| IHC Xception | Normal/ benign | 0.93 | 0.79 | 0.77 | 0.84 [0.69–0.96] | 0.90 [0.76–1.00] | 14 |
| | ADH/LG-DCIS | 0.25 | 1.00 | 1.00 | 0.40 [0.00–1.00] | 0.94 [0.80–1.00] | 4 |
| | HG-DCIS | 0.86 | 0.92 | 0.75 | 0.80 [0.55–1.00] | 0.99 [0.95–1.00] | 7 |
| | IDC | 0.63 | 0.92 | 0.71 | 0.67 [0.32–0.91] | 0.85 [0.69–0.98] | 8 |

**Supplementary Table S5. Patient-level per-class performance on GWU test cohort**

Patient-level per-class performance on the GWU external test cohort. Patient-level predictions were obtained by mean-probability pooling across all tiles from each patient. Per-class sensitivity, specificity, precision, F1 score, and AUC are reported with 95% confidence intervals estimated by bootstrap resampling (n = 500). Sensitivity, specificity, and AUC are prevalence-independent measures, whereas precision and F1 score are prevalence-dependent and should be interpreted in light of class support.

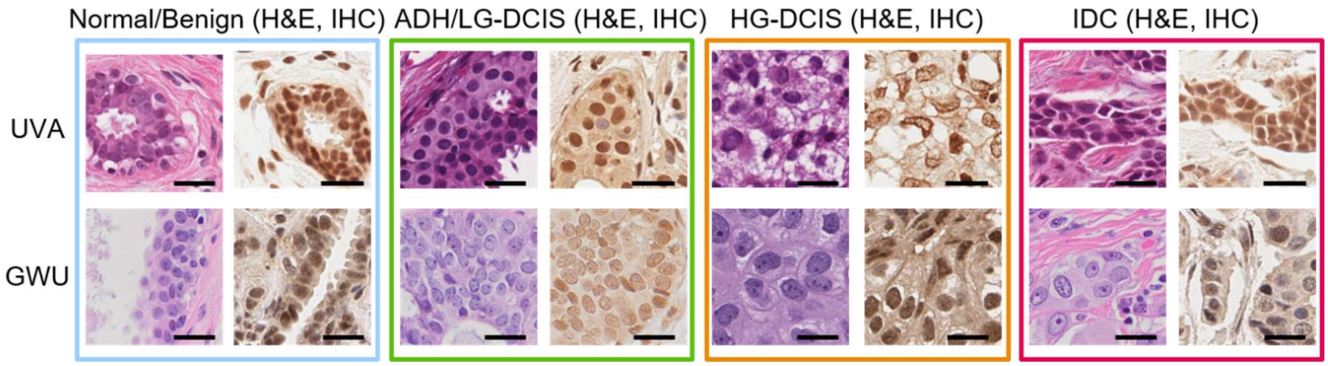


**Supplementary Figure S1. Representative 299×299 input tiles for each diagnostic group.** For each of the four ordered diagnostic groups, normal/benign (blue box), ADH/low-grade DCIS (ADH/LG-DCIS; green box), high-grade DCIS (HG-DCIS; orange box), and invasive ductal carcinoma (IDC; red box), representative tiles from the UVA (top row) and GWU (bottom row) cohorts are shown, with paired H&E (left) and TRPV4 IHC (right) tiles for each cohort. Tiles illustrate the local morphology and staining patterns used as input to the convolutional neural networks, at the same resolution and field of view as during model training and evaluation. Examples are intended to highlight typical appearances within each group rather than to capture the full spectrum of histologic variability. Scale bar, 40 μm.

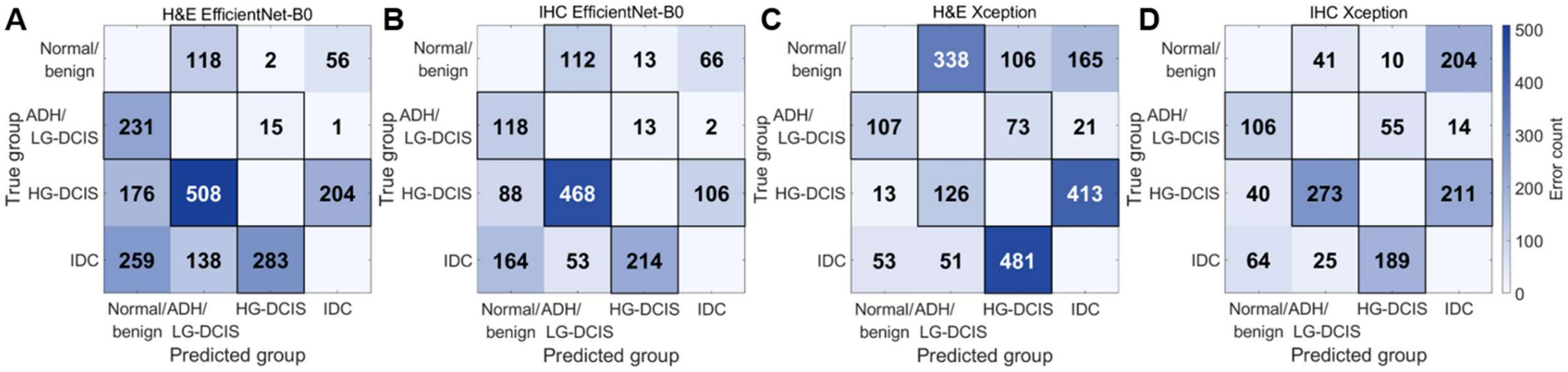


**Supplementary Figure S2. Tile-level error structure on the GWU external test cohort.**

Heatmaps show misclassified tiles (diagonal entries set to zero) for the four models evaluated on the GWU test set. **(A)** H&E EfficientNet-B0; **(B)** TRPV4 IHC EfficientNet-B0; **(C)** H&E Xception; **(D)** TRPV4 IHC Xception. Color intensity encodes the error count in each true-predicted class cell; gray boxed cells highlight adjacent-grade errors (normal/benign–ADH/LG-DCIS, ADH/LG-DCIS–HG-DCIS, HG-DCIS–IDC). For panels **A–D**, adjacent-grade errors account for 68.3% (1359/1991), 72.8% (1031/1417), 79.0% (1538/1947), and 71.0% (875/1232) of all misclassified tiles, respectively.
